# Possible impact of national responses to the COVID pandemic on medal tallies at the Paris 2024 Olympics

**DOI:** 10.1101/2024.08.23.24312521

**Authors:** John W. Orchard, Nathan Luies, Robert J. Buckley, Adam Castricum

## Abstract

**Introduction:** Western Pacific nations have experienced lower excess mortality compared to rest of the world since 2020 and recently performed exceptionally well on the medal tally at the 2024 Paris Olympics. This study aimed to analyse any possible connection between these factors.

**Methods:** The top performing 18 nations from 2012, 2016 and 2020 Olympics (after Russia and Ukraine were excluded) had their relative Gold medals, total medals and medal points (Gold =3, Silver=2, Bronze=1) for Paris 2024 analysed using a backward stepwise linear regression model. Initial input factors included previous medal tallies, home city advantage, time zone effects, national excess deaths 2020-2023, average GDP growth 2020-2023 and number of country signatories to the Great Barrington Declaration (GBD), with factors >P=0.10 removed sequentially.

**Results:** Total medals were best predicted by previous total medals (t=21.0, P<0.001) and home city advantage (t=4.1, P<0.001). Gold medals were best predicted by previous Gold medals (t=10.3, P<0.001), low national excess deaths (t=-3.2, P<0.007) and low signatories to the GBD (t=-2.2, P<0.05). Medal points were best predicted by previous medal points (t=18.1, P<0.001), home city advantage (t=3.2, P<0.007) and low national excess deaths (t=-1.8, P<0.09).

**Discussion:** The Western Pacific countries with a COVID-cautious national perspective (Australia, China, Japan, New Zealand, South Korea) tended to win more Gold medals than expected in Paris, compared to countries with a COVID-stoical national perspective (e.g. Great Britain, United States) which won fewer Golds than expected. This suggests that a COVID-cautious mentality may have contributed to better performance than a COVID-stoical approach. It is unclear whether any mechanism was physiological (less infectious disease impact before or during the Olympics) or psychological. If this effect existed for Golds, it did not appear to have any effect on Silver and Bronze medals.

**Graphical abstract:** 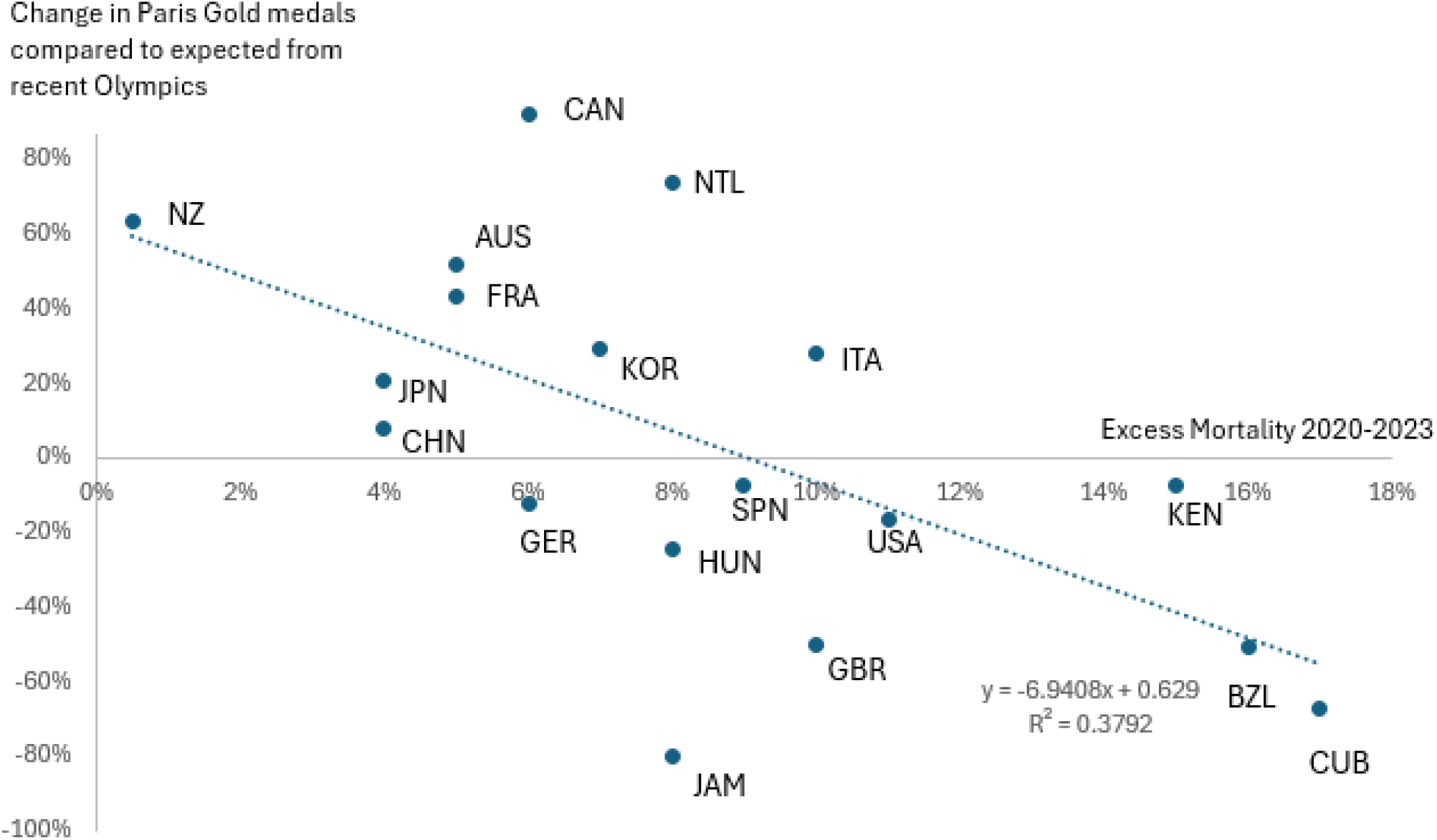

The four countries with higher than 10% Excess mortality over the time period 2020-2023 all won fewer Gold medals than expected (based on Gold medals won in the three previous Olympiads). The five countries with lower than 6% Excess mortality over the time period 2020-2023 all won more Gold medals than expected in Paris. The relationship between Excess mortality and change in Gold medals was negative and moderately strong. In the linear regression for prediction of Gold medals in Paris, Excess mortality was a highly significant predictor (t=-3.2, P<0.007). The exact mechanism of this relationship (physiological via reduced infection or psychological or via unassessed confounders) is unclear, but it can be confidently stated that in the countries which had tighter pandemic restrictions in 2020-21 and lower excess mortality, athletes have not suffered any “immunity debt” relative to the rest of the world.

## Introduction

### The Paris Olympics

Paris 2024 was the successful completion of the second summer Olympic Games since the onset of the COVID-19 pandemic and a reminder of the world-unifying nature of the Olympics. Somewhat ironically, Tokyo 2020 (postponed until 2021) was officially conducted during “the pandemic” and because of biosecurity precautions there were hardly any COVID (and other illness) cases.^1^ Paris 2024 was conducted “after the pandemic had officially ended” yet (according to media reports) the Olympics were rife with COVID and other illness.^2^ As a microcosm of the world, COVID the disease flourished in Paris when the infrastructure had attempted to return to “normal” operations.

For the Western Pacific region, the Paris Olympics were a source of regional pride, as the major nations in this area exceeded performance expectations. This had also happened at the Tokyo Olympics, but being a “home” Games regionally, success for these nations was more anticipated.^3^ There are five nations from the Western Pacific region that tend to do particularly well at the Olympics: Australia, China, Japan, New Zealand and South Korea and all five excelled in Paris, particularly with respect to Gold medals.

### The COVID pandemic response in the Western Pacific

The Olympics “Big 5” of the Western Pacific are countries of varying politics and demographics. All are relatively wealthy, which is a prerequisite for Olympic success in all regions.^4 5 6^ Australia and New Zealand are very similar nations, politically and demographically, as are Japan and South Korea, but these two sibling groups differ from each other as do all four of these nations from China. The five nations are most bound by being regional neighbours. A factor which is somewhat overlooked in 2024, but which was very prominent in 2021, is that the Western Pacific region nations would have won the retrospective “Gold medals” (were there such a thing) for COVID response during 2020 and 2021. Excess mortality veered into negative territory for some of the Western Pacific nations in 2021 (meaning there were fewer deaths than normal in the midst of a worldwide pandemic), when death rates were extremely high in most other places in the world.^7 8^ The successful outcomes for the Western Pacific nations were achieved by a combination of border closures, quarantine, lockdowns and mask mandates.^8-10^ In the case of East Asian countries, their response to the SARS-2 pandemic was perhaps driven by the lessons learned from the original SARS-1 outbreaks in the 2000s. For the Oceanic nations of Australia and New Zealand, there was less cultural preparedness but these two countries took full advantage of their geographic isolation to close their borders in early 2020.^7^

These two seemingly unrelated regional observations - Paris Olympic Gold medal success and effective containment of COVID in the pandemic phase raise an intriguing question: could they be in any way related?

This paper looks to rapidly-analyse Paris Olympic medal results to investigate any potential correlation between COVID outcomes and Olympic performance. It builds on the pre-existing literature around prediction of success in the summer Olympics, for which there are abundant previous analyses. ^4 5^

A recent point of contention is whether Gold medals are more important than total medals won.^11^ With this contention, we decided to analyse both Gold medals, total medals, and a hybrid “medal points haul” using a system of 3 points for a Gold medal, 2 points for a Silver and 1 point for a Bronze medal. A conclusion of most published studies is that national success in recent previous Olympic Games is the best predictor of success in the future. ^4, 5^ We focussed on the 20 leading nations of recent Summer Olympics (to eliminate the high random variation amongst the less successful countries). Our analysis started with the Major Outcome variables (x axis) of Paris Golds, total medals and medal points won in the Paris Olympics. Our expected major predictor variables were the same medal metrics combined for 2012 London, 2016 Rio and 2020 Tokyo Olympics by each of the same nations. The additional predictor variables we considered were: (1) host nation advantage; ^3 4^ (2) time zone effects ^3^; (3) Excess Deaths observed during the COVID pandemic,^7^ which we postulated to perhaps be negatively correlated with performance, (4) GDP growth % from 2020 to 2023,^12^ which was postulated to perhaps be positively correlated with performance. After initial exploratory analysis that did reveal Excess Deaths negatively correlating with Gold medals won in Paris, we chose to add another potential predictor variable: (5) National signatories for the Great Barrington Declaration (GBD).^13^ The GBD was an open letter authored early in the pandemic (October 2020) which claimed that public health authorities had over-reached during the pandemic response to the detriment of young adults and children. It claimed that the effects of being too cautious regarding COVID (with border closures, lockdowns, school closures, mask and subsequently vaccine mandates) would have a greater negative effect on the young populations than the negative effects of greater COVID spread itself (which they argued were trivial in young people). We postulated that countries with high numbers of GBD signatories would reflect psychologically those countries with the greatest contrast to the Western pacific nations in terms of preferred COVID response (stoical vs cautious).

Given the global impact of the COVID pandemic, understanding whether successful management strategies have extended benefits beyond public health to influence international sporting competition is crucial. In particular, the Olympics are a competition between a nation’s “young adults”, who were the population demographic claimed to have been most harmed by authors of the Great Barrington Declaration. This study aims to determine if there is a quantifiable relationship between a nation’s pandemic response and its Olympic performance.

## Methods

### Medal counts and selection of nations to analyse

Medal counts for the 2012, 2016, 2020 and 2024 Summer Olympics (Golds, total medals and according to the 3-2-1 system) and were sourced from Wikipedia/IOC websites on August 12, 2024 at the conclusion of the Paris Olympiad. Using the points system and combining 2012-2020 Olympiads, the previous top 20 nations were clearly defined with all these nations performing well at all 3 previous Olympiads. Because of their massive reductions in team sizes for Paris 2024, Russia and Ukraine were excluded from the analysis leaving 18 teams (Table 1).

**Table 1.** Overall outcome variables and potential predictor variables for the 18 countries in the analysis.

| Country | Outcome variables<br>(shaded mauve) |  |  | Predictor variables (primary shaded Orange, possible shaded Blue/White) |  |  |  |  |  |  |  |
| --- | --- | --- | --- | --- | --- | --- | --- | --- | --- | --- | --- |
|  | Paris Gold | Paris medals | Paris haul | Prev Gold | Previous medals | Prev haul | Excess mortality | GBD sigs | Home advt | Time Zone | GDP 20-23 |
| United States | 40 | 126 | 250 | 133 | 338 | 708 | 11% | 100000 | 0 | -2 | 2.02% |
| China | 40 | 91 | 198 | 103 | 251 | 538 | 4% | 1000 | 0 | -1 | 4.73% |
| Japan | 20 | 45 | 97 | 46 | 137 | 265 | 4% | 2000 | -0.8 | -1 | 0.32% |
| Australia | 18 | 53 | 108 | 33 | 110 | 209 | 5% | 12000 | 0 | -1 | 2.32% |
| France | 16 | 64 | 122 | 31 | 110 | 213 | 5% | 10000 | +3 | 0 | 0.55% |
| Netherlands | 15 | 34 | 71 | 24 | 75 | 148 | 8% | 10000 | 0 | 0 | 1.68% |
| Great Britain | 14 | 65 | 115 | 78 | 196 | 413 | 10% | 100000 | -1.2 | 0 | 0.89% |
| South Korea | 13 | 32 | 67 | 28 | 72 | 144 | 7% | 1000 | 0 | -1 | 1.89% |
| Germany | 12 | 33 | 70 | 38 | 123 | 240 | 6% | 100000 | 0 | 0 | 0.21% |
| Italy | 12 | 40 | 77 | 26 | 96 | 179 | 10% | 10000 | 0 | 0 | 1.06% |
| New Zealand | 10 | 20 | 47 | 17 | 51 | 102 | 1% | 2000 | 0 | -1 | 2.21% |
| Canada | 9 | 27 | 52 | 13 | 64 | 106 | 6% | 60000 | 0 | -2 | 1.29% |
| Hungary | 6 | 19 | 38 | 22 | 53 | 111 | 8% | 1000 | 0 | 0 | 2.37% |
| Spain | 5 | 18 | 32 | 15 | 54 | 91 | 9% | 10000 | 0 | 0 | 0.88% |
| Kenya | 4 | 11 | 21 | 12 | 36 | 74 | 15% | 1000 | 0 | -1 | 4.06% |
| Brazil | 3 | 20 | 33 | 17 | 57 | 108 | 16% | 3000 | -1.0 | -2 | 1.85% |
| Cuba | 2 | 9 | 14 | 17 | 41 | 83 | 17% | 500 | 0 | -2 | 0.00% |
| Jamaica | 1 | 6 | 11 | 14 | 33 | 70 | 8% | 500 | 0 | -2 | -0.03% |

### Home country advantage

A home country advantage metric was calculated as follows:

+3 points for France (as host of the Paris Olympics);

-1.2 points for Great Britain, -1 point for Brazil and -0.8 points for Japan (loss of hosting benefit from the predictor period), which was to account for anticipated staggered loss of medals due to previous hosting benefit having worn off; ^3^

Zero was awarded for the hosting effect for the other 14 nations;

### Time zone effects

A Time Zone (TZ) effect variable was calculated as follows (based on the work of Jasper et al.)^3^:

-1 time zone points for Kenya, Australia, China, Japan, New Zealand, South Korea (for Westward travel to Europe);

-2 time zone points for Brazil, Canada, Cuba, Jamaica and United States (for Eastward travel to Europe);

0 points were awarded for the 7 European nations who were competing in approximately their home time zone and in their home region.

### Excess mortality resulting from the COVID pandemic

National excess mortality to the end of 2023 which took into account differences in COVID-related national mortality during the pandemic time period (from 2020 onwards).^7^ Where December 2023 figures were unavailable the closest other time point was used as an approximation. As specific figures for Kenya were unavailable, South Africa was used as the closest country approximation. As figures for China were unavailable, Japan was used as the closest country approximation (of the closest Asian nations to China, only Japan had data up until December 2023 available).

### GDP growth during the pandemic period

Average national GDP growth from 2020-2023 ^12^ was obtained also from Our World in Data on the same date (12/8/24). For some countries we used an average of 4 years (2020-2023) and others we used 3 years if the 2023 data was not available. Cuba was given a null value of 0% growth (neither positive nor negative) as GDP data was unavailable.

### Great Barrington Declaration signatories

We estimated number of signatories from each country of the world to the Great Barrington Declaration on 12/8/24, using the colour signature map provided on their own website.^13^

### Technique for linear regression and other statistical analysis

Linear Regression with a backward stepwise methodology (P > 0.10 threshold to remove), was conducted in Microsoft Excel. Linear regression has been the primary statistical technique used for prior predictions of Olympic medals.^4, 6^

A scatter correlation graph was also prepared in Excel with trendline incorporated to show the raw relationship between change in Paris Gold medal count (compared to historical) and Excess pandemic deaths for each of the 18 nations

### Ethical considerations

No ethical clearance was required for this study as it used publicly-available data only (for countries) and did not involve any individuals. We have declined to name any individuals in this analysis as it was not necessary (although some of the references may name individuals).

## Results

### Input variable summary

The top 18 nations – after removal of Ukraine and Russia - according to medal points haul from 2012, 2016 and 2020 Olympics are listed in Table 1. The 3 main outcome variables (2024 Gold medals, 2024 total medals, and 2024 medal points haul) are listed along with the expected major predictor variable (previous results) and possible additional predictor variables.

### Linear regression results

The best predictor by far for medals in Paris was the corresponding variable for London, Rio and Tokyo Olympics combined. The P-values for previous medals predicting Paris medals were all P<0.0001 for total medals (Table 2), Gold medals (Table 3) and medal points hauls (Table 4). No other variable consistently appeared in all 3 linear regression equations, although home advantage (positive effect) and excess mortality (2020-2023), which had a negative effect meaning that nations which had higher excess mortality won fewer medals, appeared in two out of the three regression equations (at P<0.10 or better). The only other variable that was included in one out of the three regression equations was Great Barrington Declaration signatures, which was associated with fewer Gold medals only (P<0.05). GDP change (from 2020-2023) and the time zone effect variable were not included in any final regression equations.

**Table 2.** Linear regression analysis to predict Total medals at Paris (R^2^ = 0.967)

| *Included variables* | *Coefficients* | *Standard Error* | *t Stat* | *P-value* |
| --- | --- | --- | --- | --- |
| Intercept | 0.509 | 2.336 | 0.22 | 0.8304 |
| **Previous medals** | 0.371 | 0.018 | 20.95 | 0.0000 |
| **Home advantage** | 7.006 | 1.718 | 4.08 | 0.0010 |
| Non-included variables |  |  | T when removed | P when removed |
| GBD signatures |  |  | -1.21 | 0.243 |
| GDP change |  |  | 0.59 | 0.568 |
| Excess mortality |  |  | -0.35 | 0.730 |
| Time zone effects |  |  | 0.02 | 0.986 |

**Table 3.**
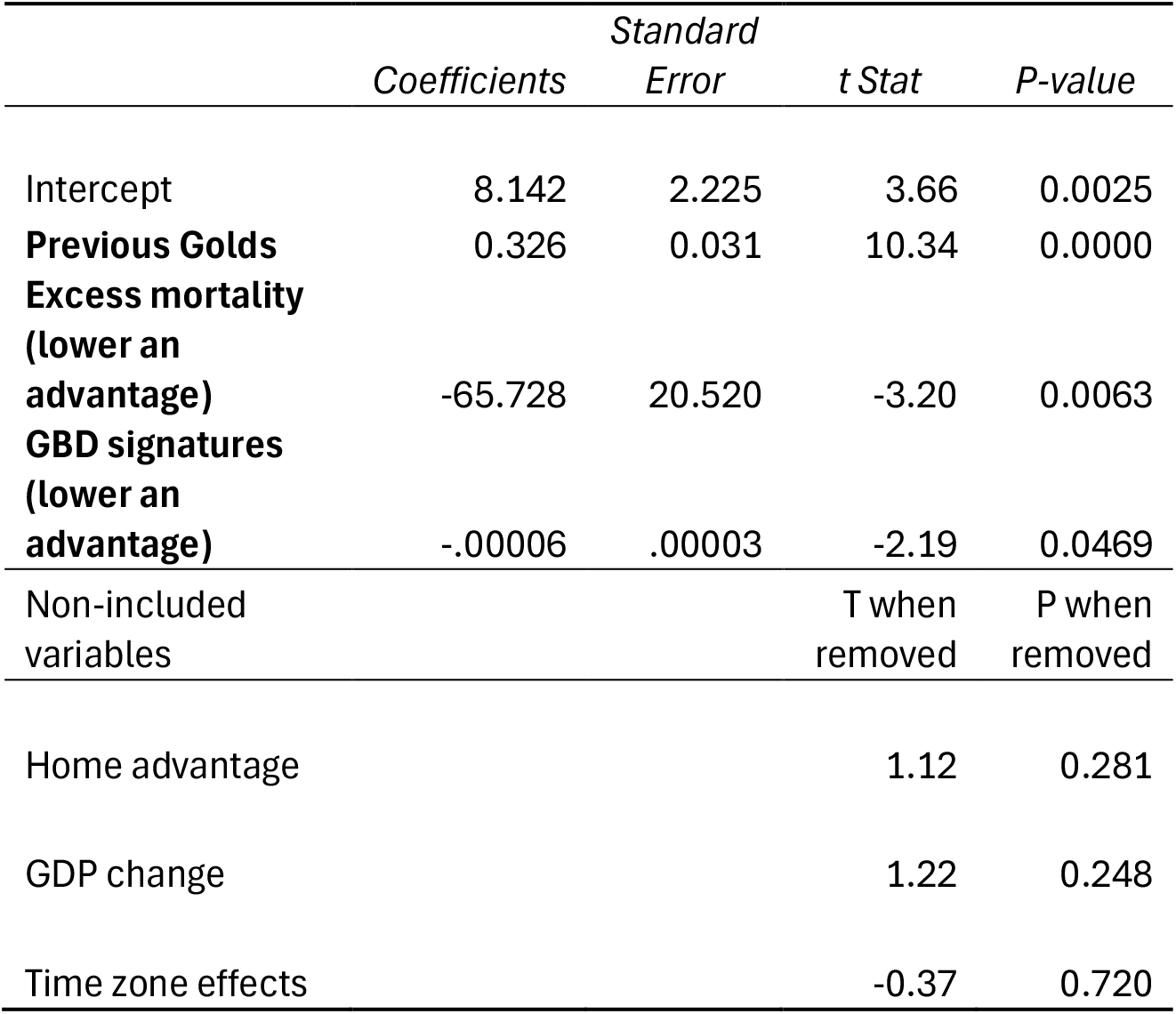
Linear regression analysis to predict Gold medals at Paris (R^2^ = 0.911)

**Table 4.**
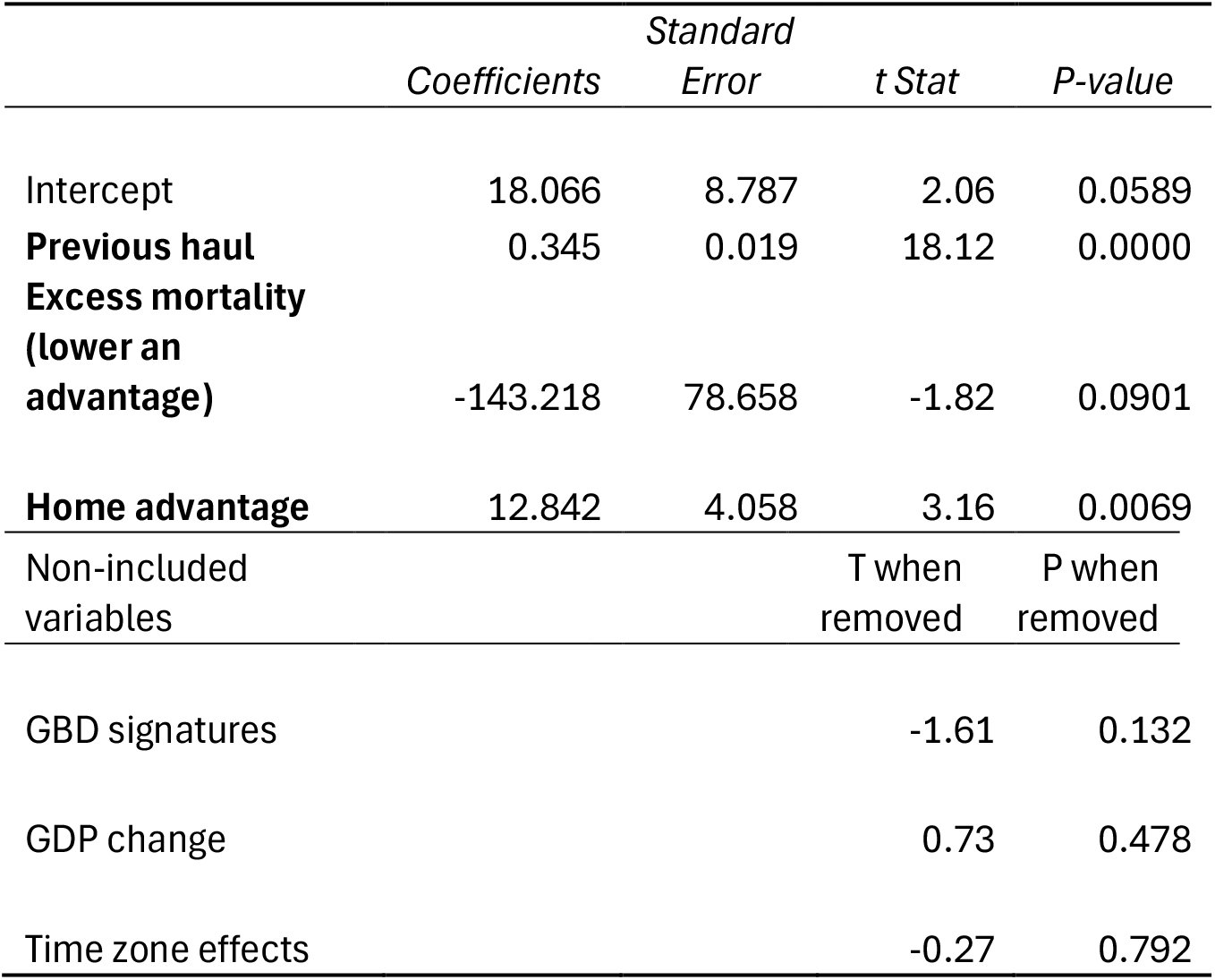
Linear regression analysis to predict medal haul (G3, S2, B1) at Paris (R^2^ = 0.962)

### Effect of excess mortality

The relative advantage of low Excess mortality (2020-2023) was not a predictor of total medals (Table 2) but was a strong predictor of Gold medals in Paris (Table 3, P=0.006) and a weak predictor of overall medal haul in Paris (Table 4). Countries with lower excess mortality tended to win more Gold medals than expected and vice versa. When excess mortality was plotted against change in Gold medals (from previous Olympiads to Paris) there was a moderately strong relationship (Figure 1).

**Figure 1.**
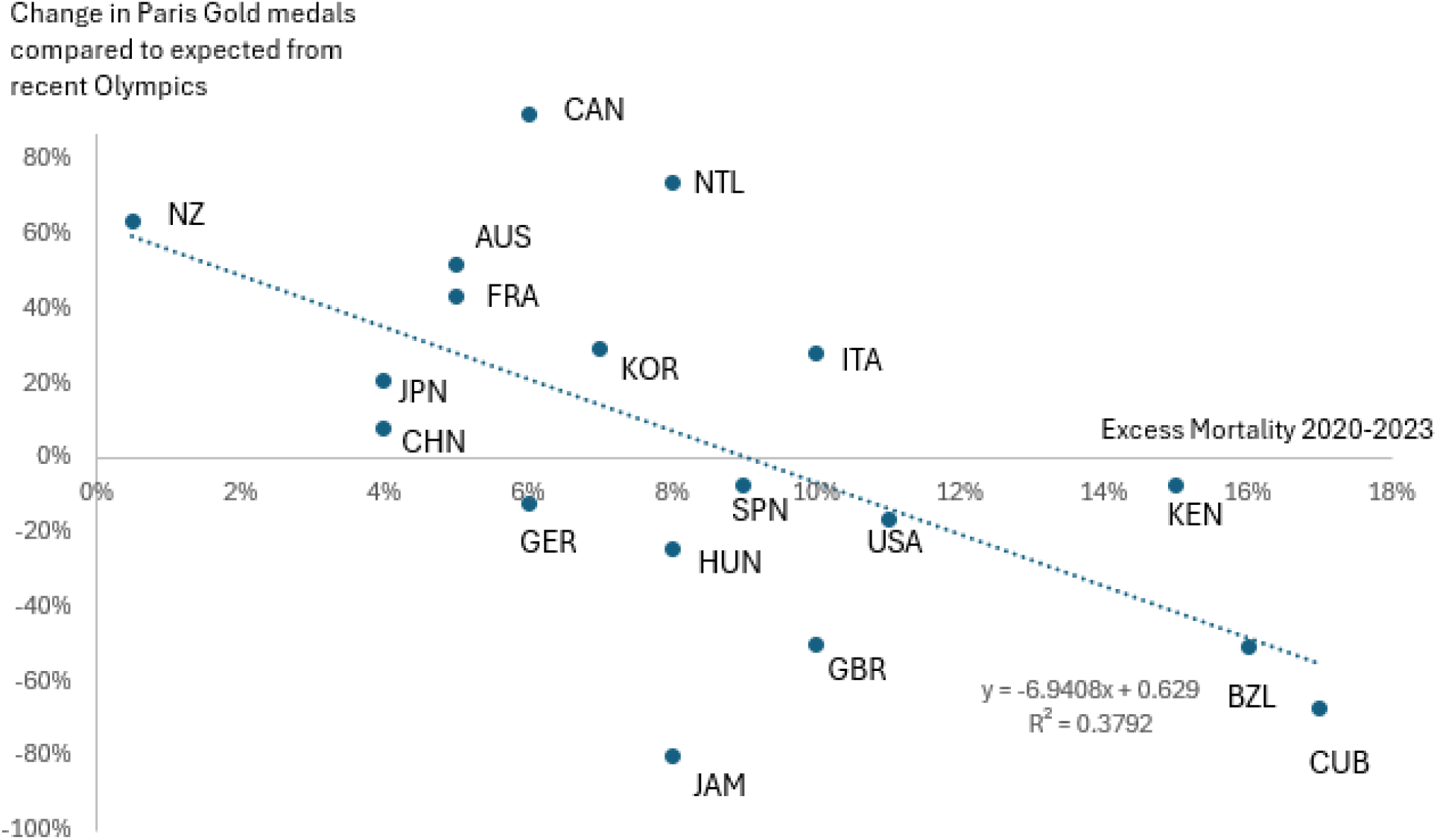
Moderately strong correlation (R^2^ = 0.379) between change in Gold medal haul in Paris (y-axis) and Excess mortality 2020-2023 (x-axis).

### Home city advantage

There was a strong relationship between home city advantage and total medals won in Paris (P<0.001), but only a weak relationship between home city advantage and both Gold medals won and medal haul points.

### Great Barrington Declaration signatures

The number of GBD signatures was significantly associated (P<0.05) in a negative relationship with Gold medals won in Paris. That is, countries who provided a high number of signatories to the Great Barrington Declaration won fewer Gold medals than expected in Paris. However there was only a weak (negative) relationship between GBD signatures and total medals and medal points haul.

### GDP change and time zone effects

Neither change in GDP (2020-2023) for countries nor time zone effects were significantly associated with medals won in Paris.

## Discussion

### Paris Gold medals were associated with pandemic response variables

The regression analysis for total medals at the Paris Olympics was relatively unremarkable, with previous recent total medals and home advantage being the only significant predictors.

However, when the focus was on Gold medals alone (or a hybrid formula of medal haul where Golds were considered more important than minor medals) then the regression analysis results were very different. Previous Gold medals again was by far the best predictor of Paris Gold medals, which was unsurprising. However, home advantage was relatively less correlated and Excess mortality become a weak predictor of medal haul and a very strong (negative) predictor of Paris Gold medals (t = -3.2, P = 0.0063). All of the Western Pacific group of nations (Australia, China, Japan, New Zealand and South Korea) won more Gold medals in the 2024 Olympics than expected from previous Games, and they also generally had lower excess mortality than other countries. We consider these Western Pacific nations to have had a “COVID cautious” national psyche, which helped them avoid excess deaths particularly prior to vaccination rollouts. The countries with a “COVID stoical” national psyche were those more likely to have challenged public health measures and convince their governments to “live with/alongside the COVID virus” and to abandon containment measures in young people. These countries tended to have high signatories to the Great Barrington Declaration.

### Possible confounders

The most interesting discussion point is whether the correlation between low excess deaths and Olympic success suggests any causative relationship; and alternatively, how many unassessed confounders might be involved.

At a starting point, we consider it very unlikely that there were many potential Olympians who died in the period 2021-2024 and were therefore removed from the population of possible competitors for their countries. An exception to this statement might be in the war zones (hence the removal of Ukraine and Russia from the analysis); outside war zones in terms of high-profile deaths there was only a Kenyan marathon runner who tragically died in a car accident aged 24. Studies of sudden cardiac death in the young over the pandemic period has not yet found a significant increase in young athletes,^14^ despite the clear increase in deaths affecting older age groups. It is very unlikely that nations with high excess deaths literally had athletes die and therefore be unable to represent their country.

It is more plausible that Excess mortality served as a surrogate for the overall metrics of the pandemic, including the total number of cases and their severity. We chose Excess mortality as our preferred predictor variable because it has been noted that actual recorded COVID cases was unreliable, often influenced by the varying levels of testing conducted.

The most important potential confounder that we were able to remove as not responsible was economic growth (change in GDP), which did not have any significant predictive effect.

Other (albeit smaller) potential confounders might include the distribution of new Olympic events, and whether this favoured certain nations. This is likely to have only a tiny effect in that the number of medals allocated to new events is very small, and because the host nation has an influence on new events it is also likely to be accounted somewhat by the home advantage effect. Another confounder was the removal of Russia - traditionally in the top 3-4 nations in the medal count - from the pool of competitors. It is challenging to estimate whether the absence of Russia as a competitor benefitted Western Pacific nations over European nations. The Soviet Union previously boycotted the Los Angeles Olympics in 1984 which was seen to be of primary benefit to the USA, but it is problematic and fraught to try to estimate which medals Russia might have won (and which countries they would therefore have displaced) if it was at full strength at Paris 2024.

Some of the other confounders would have been accounted for by the previous medal counts (e.g. population size and density, country weather patterns, existing health systems) and have been analysed in previous similar papers. ^4 5 6^

### Physical versus psychological effects of infectious diseases

A further debate is to what extent the direct effects of COVID itself might explain changes in Olympic performance, as opposed to any lingering psychological effects of the pandemic restrictions - this was our rationale for including the Great Barrington Declaration signatures in the analysis. Additionally there has also been debate about whether young people from the countries with the strictest measures in 2020 and 2021 would have possibly suffered “immunity debt” due to reduced exposure to circulating respiratory viruses (not just COVID but all types).^15^ Alternatively it is possible that young people in less restrictive countries may have suffered “immune theft or damage” from having contracted COVID more frequently.^16^

Our results conclusively reject any notion that athletes from Western Pacific nations have suffered harm from “immunity debt” as the analysis shows a beneficial correlation between less exposure to COVID and other respiratory viruses and greater Olympic performance. However the evidence is less conclusive regarding whether athletes from the Americas or Europe experienced immune damage that lead to performance disadvantage.

The direct and delayed effects of COVID on athlete performance appear to be bioplausible. COVID has been associated with decreases in population IQ,^17^ increased numbers of traffic accidents,^18^ increased risk of diabetes, reduced cardiopulmonary fitness^19^ and other diseases along with reductions in life expectancy.^20^ The percentage of athletes suffering “long COVID” after infection is around 4%,^21^ far lower than for the general population.^22^ Long COVID and health effects are worse in the unvaccinated population.^23^ It can be confidently stated that the majority of athletes in Australia, China, Japan, Korea and New Zealand would have been vaccinated before their first COVID infection. In contrast, this is less likely for athletes from other countries in the analysis, as COVID was circulating at far higher rates in these countries prior to the vaccine rollout. ^21^ If this association alone led to lower rates of long COVID in the Western Pacific nations, this could explain a relative improvement in athletic performance.

Sensible recommendations to avoid COVID and other respiratory illnesses in the Olympic Village in Paris have been recently published.^24^ It stands to reason that the COVID-cautious athletes from the Western Pacific region may have been more adherent to preventive measures than the athletes from more COVID-stoical regions, and hence were less affected by COVID (and potentially other illnesses) during the Olympics itself. There were multiple media reports of Silver and Bronze medallists revealing afterwards that they had been suffering from COVID during the Games.^2^ Even though COVID might be now a more trivial disease in healthy young athletes, it could easily represent the difference between Gold and a minor medal.

Perhaps the impact was more psychological than physical (which would likely have affected total medal counts). The three countries who provided the most signatories to the Great Barrington Declaration^13^ - the United Kingdom, United States and Germany all had reasonable (close to expected) total medals won in Paris but had drops in the number of Gold medals won (compared to expected). The battle to win Gold over Silver is sometimes won in the mind rather than with the body. A COVID outbreak in the Olympic village might be challenging but perhaps not intimidating to a COVID-cautious athlete from a Western Pacific nation, who may have had expectations of this happening and for which they were prepared (with booster vaccination, masks, anti-viral medications, isolations plans etc.). The same outbreak might be more intimidating to a COVID-stoical athlete from a country with a national psyche that “COVID does not really affect healthy young people” when the immediate evidence they were observing in the Olympic Village was that it can. There is also some research linking the physical to psychological showing that COVID infections result in reduced wellness and mental well being.^25^

## Conclusion

In conclusion, this paper raises an interesting association between stricter pandemic restrictions in the Western Pacific nations, lower resultant excess mortality and subsequent Olympic success in the Paris Olympics. Whether this is a coincidental association, causative in a physical or psychological sense, or is best explained by unknown confounders remains unanswered. However, it can be confidently stated that athletes from Western Pacific nations suffered no ill effects from the strict pandemic response in their countries in 2020-2021, relative to the rest of the world. Countries should all, of course, continue to try to contain COVID simply to try to reduce rates of excess mortality evident since the start of the pandemic, for its own sake. Citizens (whether old or young) dying earlier than they would have otherwise is a more important issue than Olympic medals. This study suggests that there is no trade-off required, providing some indirect evidence that healthy young people also may benefit from a national outlook that tries to reduce the rates of circulating COVID (and other infectious disease).

## Data Availability

A supplemental Excel file containing all data used for the analysis is included as an attachment

## Notes

### Competing Interest Statement

The authors have declared no competing interest.

### Funding Statement

This study did not receive any funding

